# Maximum Wall Thickness and Papillary Muscle Hypertrophy as Complementary Cardiac Biomarkers in Fabry Disease

**DOI:** 10.64898/2026.05.06.26352512

**Authors:** Magdalena Schüttler, Jannis Witte, Peter Nordbeck, Magnus Schindehütte, Markus Ankenbrand

## Abstract

**Background:** Fabry disease (fd) is a rare and severe disease affecting multiple organ systems. However, its non-specific and heterogeneous presentation poses a critical challenge for early diagnosis, often delaying necessary treatment. In recent years, imaging-based biomarkers have been increasingly proposed to improve the understanding of fd and aid its diagnosis. This study presents a comprehensive comparative analysis of several previously proposed imaging-based cardiac biomarkers to assess their potential for diagnostic use.

**Methods:** We have developed a fully automated image analysis pipeline for quantifying cardiac metrics based on short-axis cine cmr data available on the UK Biobank.

**Results:** Based on the UK Biobank cohort, our analyses confirm the diagnostic relevance of the maximum myocardial wall thickness, a metric that mimics the current clinical practice for diagnosing left ventricular hypertrophy. Initial evidence also suggests that the PM/LV ratio, which measures the papillary muscle hypertrophy as the ratio between the areas of the papillary muscles and the left ventricular cavity, has potential prognostic relevance.

**Conclusion:** This study contributes towards a better understanding of the cardiac presentation of FD, which may support future research in improving the diagnostic process. Additionally, our analysis pipeline can serve as a valuable basis for additional data analysis of imaging-based biomarkers for fd and other diseases.

## 1 Introduction

Fabry disease (fd) is a rare lysosomal storage disorder caused by a mutation of the *GLA* gene. fd presents itself with a highly variable clinical presentation and mostly non-specific symptoms, impeding early and accurate diagnosis and delaying therapeutic intervention. However, lack of treatment often entails severe consequences, including cardiac and renal failure, as well as neurological impairment. Meanwhile, the two main diagnostic methods, genetic sequencing and enzyme activity assays, are critically limited for female patients and patients carrying gene variants of unknown pathogenicity. This results in a substantial number of overlooked cases or misdiagnoses. Thus, the improvement of diagnostic approaches is currently a major focus of fd research [1, 2].

Cardiac manifestations are common in fd and frequently present as left ventricular hypertrophy (lvh). It is typically considered a late marker in FD, appearing only when other cardiac damage is already present. Nevertheless, idiopathic lvh is often the first indicator prompting consideration of fd [1–3].

Left ventricular papillary muscle hypertrophy (pmh) is also prevalent among fd patients [4]. Papillary muscles are small muscles located within the left ventricle. Their main function is to support the mitral valve throughout the cardiac cycle. Classically, two papillary muscles—one anterolateral and one posteromedial muscle—are described, but a majority of people exhibit divergent morphology, often without showing any symptoms of cardiac dysfunction [5, 6]. While pmh was proposed as an earlier and more specific marker for fd several years ago [7, 8], its role remains incompletely understood [4]. Cianciulli et al. [4] presented some evidence that pmh has a predictive role for other cardiac damage, specifically LVH.

Both lvh and pmh are commonly diagnosed clinically and quantified in research with echocardiography. Cardiac magnetic resonance imaging (cmr) is also used, although less frequently [3, 9].

In this study, we aim to assess lvh and pmh as imaging-based biomarkers for diagnostic and prognostic use in FD. To this end, we perform a comprehensive, comparative analysis of multiple previously proposed cardiac metrics using cmr data provided by the UK Biobank. To quantify and assess the metrics, we have developed a fully automated image processing pipeline, designed for use on the UK Biobank Research Analysis Platform (UKB RAP). These analyses will contribute to a better understanding of cardiac involvement in fd and establish a baseline for further research on cardiac biomarkers.

## 2 Materials and Methods

### 2.1 Available Data

For this study, we are utilizing data provided by the UK Biobank. This dataset features genetic sequences and medical records of 502 150 participants, as well as multimodal medical imaging data of 83 469 participants as the time of this study. With its standardized imaging protocols and data structure, the UK Biobank is a valuable and powerful tool for research of multi-organ diseases such as FD.

For the identification of the fd cases, reliance on diagnostic records proved insufficient. Sole reliance on available hospital records would overlook late and misdiagnosed cases, as well as cases without hospital admission. However, exclusion of these sub-populations of fd cases would introduce bias towards classical and severe phenotypes. Thus, their inclusion is critical for the research of diagnostic methods. Instead, we identified fd cases based on known pathogenic and likely pathogenic variants in the *GLA* gene, collected from databases and literature. This way, we identified 92 fd patients throughout the UK Biobank dataset, 18 of whom underwent relevant short-axis cine cmr scans. This fd group comprises predominantly female participants (15 of 18) and includes three patients with an ICD-10 code E75.2, which includes fd diagnosis, listed in their medical records. Participants ranged in age from 47 to 78 years, with a mean age of 64.00 years. Observed *GLA* variants are listed in Appendix A. There are no repeat scans of fd patients available at this time.

We use short-axis cine cmr scans for all of the presented analyses. The scans within the UK Biobank project were performed using a 1.5 T MAGNETOM Aera magnetic resonance imaging (mri) scanner (Siemens Healthineers, Erlangen, Germany). The TRUFI (true fast imaging with steady-state free precession) pulse sequence (flip angle 80*^◦^*, TR 2.6 ms, TE 1.1 ms) provides high signal from blood, thereby allowing differentiation of the darker myocardium from the bright blood-filled cardiac cavity. At the same time, it enables short imaging time and high temporal resolution. Image dimensions of one exemplary scan are listed in Table 1. Additional information about the imaging protocol of the cmr images can be found on the UK Biobank Showcase or in Petersen et al. [10].

**Table 1:**
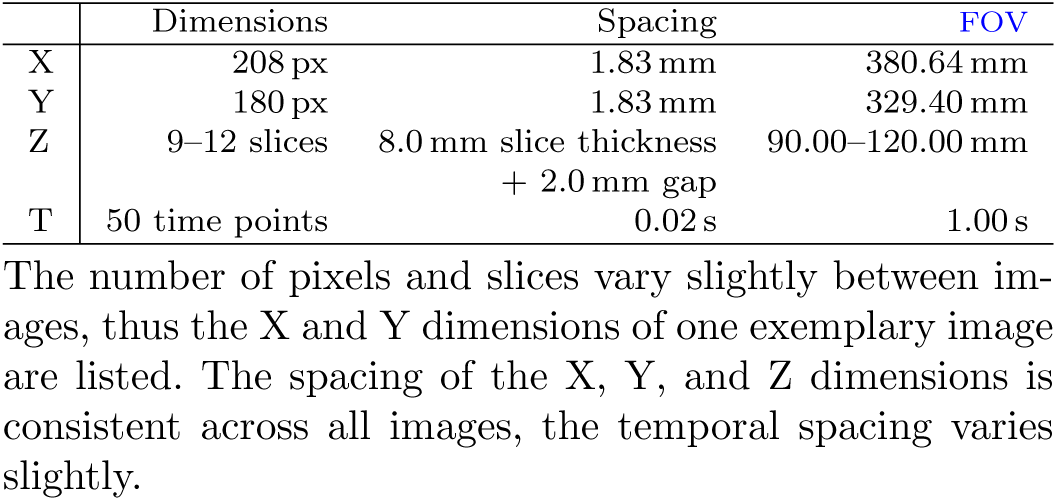
Overview of the image dimensions of the utilized short-axis cine cmr scans.

### 2.2 Utilized Software

All analyses were performed on the ukb RAP, a cloud-based platform provided for analysis of the UK Biobank data.

The image analysis pipeline is coded in Python (version 3.11.5). All utilized Python packages and respective versions are listed in the GitHub repository (linked in the Declarations Section). The segmentation and measurement of the left ventricular myocardium, which is part of the pipeline, is performed with `ukbb_cardiac` [11], a deep learning-based segmentation tool developed for and trained on the cmr imaging data available on the UK Biobank.

### 2.3 Statistical Analysis

Statistical tests were performed to compare the biomarker values between groups. For this, the normality of each group is tested using the Shapiro-Wilk test. The difference between the groups is then quantified using a two-sided Mann-Whitney U test, which does not assume normality of the data. For each group of tests, the Bonferroni-corrected level of significance *α_c_* is calculated and listed in Table 2.

**Table 2:** Bonferroni-corrected levels of significance *α_c_*.

| Group | $k$ | $\alpha_c$ |
| --- | --- | --- |
| Comparison of FD group to controls, age-correlation | 6 | 0.008 333 |
| Sex stratification | 18 | 0.002 778 |
| Pair-correlation | 16 | 0.003 125 |
| Multiple regression | 3 | 0.016 67 |

$\alpha_c$ values are calculated from the number of tests $k$ .

The effect sizes are quantified using the unitless Cohen’s *d*, allowing for comparison of effect sizes between different metrics. A *d*-value of 0.2 indicates a small effect size, a value of 0.5 a medium effect, and a value of 0.8 a large effect size, as suggested by Cohen [12]. The mean difference *MD* shares its unit with the tested metric and provides information about the observability of the difference in a clinical setting.

Receiver operating characteristic (roc) analysis provides further information about the role of a tested metric in a diagnostic setting. The roc curve visualizes the true positive rate (tpr) and false positive rate (fpr) for each possible threshold value. An area under the curve (auc) of 1 indicates perfect differentiation of the two groups based on the metric, while a random model is expected to yield an auc of 0.5.

The Pearson correlation coefficient *r* quantifies the linear correlation of two variables *x*_1_ and *x*_2_. Additionally, multiple regression models the relationship between a dependent variable and two predictor variables. Here, the ordinary least squares (ols) regression is used to fit a regression plane to the three-dimensional data. The slopes of the plane *β*_1_ and *β*_2_ and their corresponding *p*-values describe the effect of the predictor variables *x*_1_ and *x*_2_ on the response variable *y*, when correcting for the effect of the other. Notably, the *β*-values are not comparable between metrics due to the differing units.

## 3 Results

### 3.1 Cardiac Quantification Pipeline

We developed a fully automated image analysis pipeline to quantify the metrics based on short-axis cine cmr scans available on the UK Biobank. The pipeline is coded in Python and is tailored to the imaging data available on the ukb RAP, but functions platform-independently. The source code of the pipeline is available on GitHub, the link can be found in the Declarations Section.

#### 3.1.1 Data Preparation and Preprocessing

On the ukb RAP, the short-axis cine cmr scans are available as dicom files for each individual T and Z slice. The pipeline first automatically reads and sorts these slices into 4D (time-resolved 3D) image arrays using an approach largely based on work by Bai et al. [11]. The resulting image array is then narrowed down to the area of interest, the left ventricle, by cropping using a template matching approach (Figure 1a). For this, one sample image is cropped manually and used as a template. The fifth Z slice, counting from the apical side, is selected for further processing, yielding a time-resolved 2D image array. This approach is demonstrated to contain a suitable cross-section of the papillary muscles in all available cmr scans in Section 3.2.

**Figure 1:**
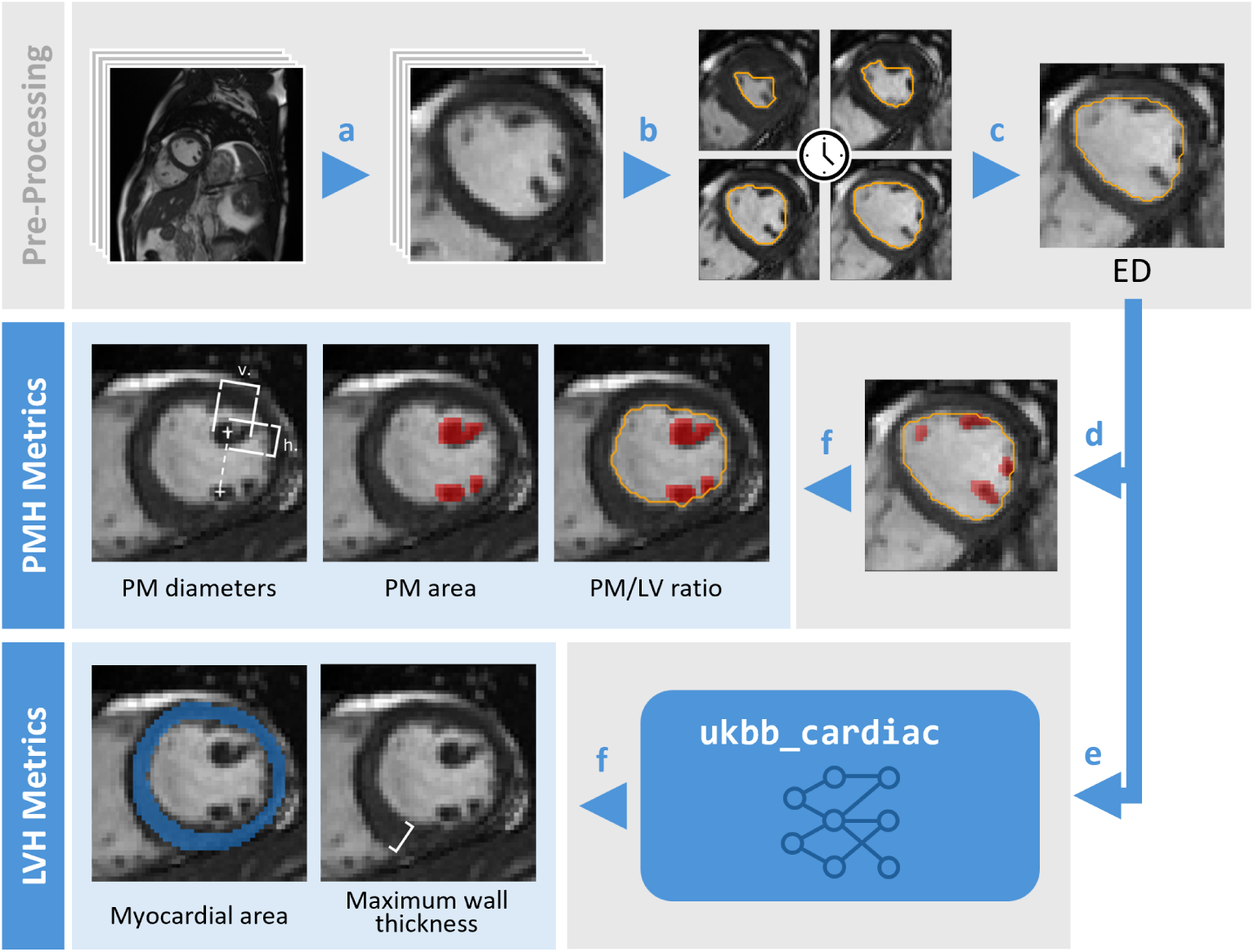
Illustration of the steps performed by the automated image analysis pipeline. cmr scans are reproduced by kind permission of UK Biobank ©.

#### 3.1.2 Segmentation

Segmentation of the blood pool (the left ventricular cavity) for each time point (Figure 1b) is achieved using Li thresholding [13] and allows for accurate definition of the cardiac phases. The selected metrics are quantified in the end-diastole (ed) time point, which is defined here as the time point where the blood pool area is at its maximum (Figure 1c). The blood pool then functions as a mask for the segmentation of the papillary muscles (PMs) (Figure 1d). For this, Otsu’s thresholding [14] is selected, as it achieves accurate area estimation as well as good differentiation of the individual PMs. The segmentation of the left ventricular myocardium is performed with `ukbb_cardiac` [11] (Figure 1e).

#### 3.1.3 Cardiac Metrics

Based on these three segmentations—the blood pool, the PMs, and the myocardium—the metrics of interest are calculated (Figure 1f).

To quantify LVH, we are investigating the following two metrics. The maximum myocardial wall thickness (mwt) mimics the typical diagnostic approach for defining lvh [9], and is measured here using the `ukbb_cardiac` module [11]. As a more comprehensive alternative, the myocardial cross-sectional area is measured, as it is more robust regarding non-concentric hypertrophy.

Cianciulli et al. [4] have proposed a way of quantifying pmh by measuring horizontal and vertical pm diameters of the two classically described PMs, yielding four diameter measurements per scan. Because of this requirement of classical morphology, we select the two PMs with the largest cross-sectional area for this metric. The PM/LV ratio was proposed by Niemann et al. [8] and further analyzed by Mattig et al. [15]. This is calculated by dividing the pm cross-sectional area by the cross-sectional area of the blood pool. Both the pm diameters and the PM/LV ratio were originally quantified manually from echocardiograms by Cianciulli et al. [4] and Niemann et al. [8], respectively. Additionally, we also evaluate the pm cross-sectional area.

In addition to the image analysis, the pipeline selects trait-matched control group participants for each fd case, as detailed in Section 3.3. Finally, the pipeline displays the segmentations for visual assessment, plots the measurements, and performs simple statistical analyses by calculating the statistical significance, the Cohen’s *d*, and the mean difference *MD* (Section 2.3).

### 3.2 Quality Control

The quality of the segmentations and measurements is assessed primarily by visual assessment. This is easily done for small and medium sample sizes that are available for this work. Figure 2 shows the segmentations of all images of the fd group. Overall, the quality of the blood pool and pm segmentations produced by the pipeline was found to be satisfactory. Adjacent PMs are generally separated well, with few exceptions that are deemed tolerable without additional intervention.

**Figure 2:**
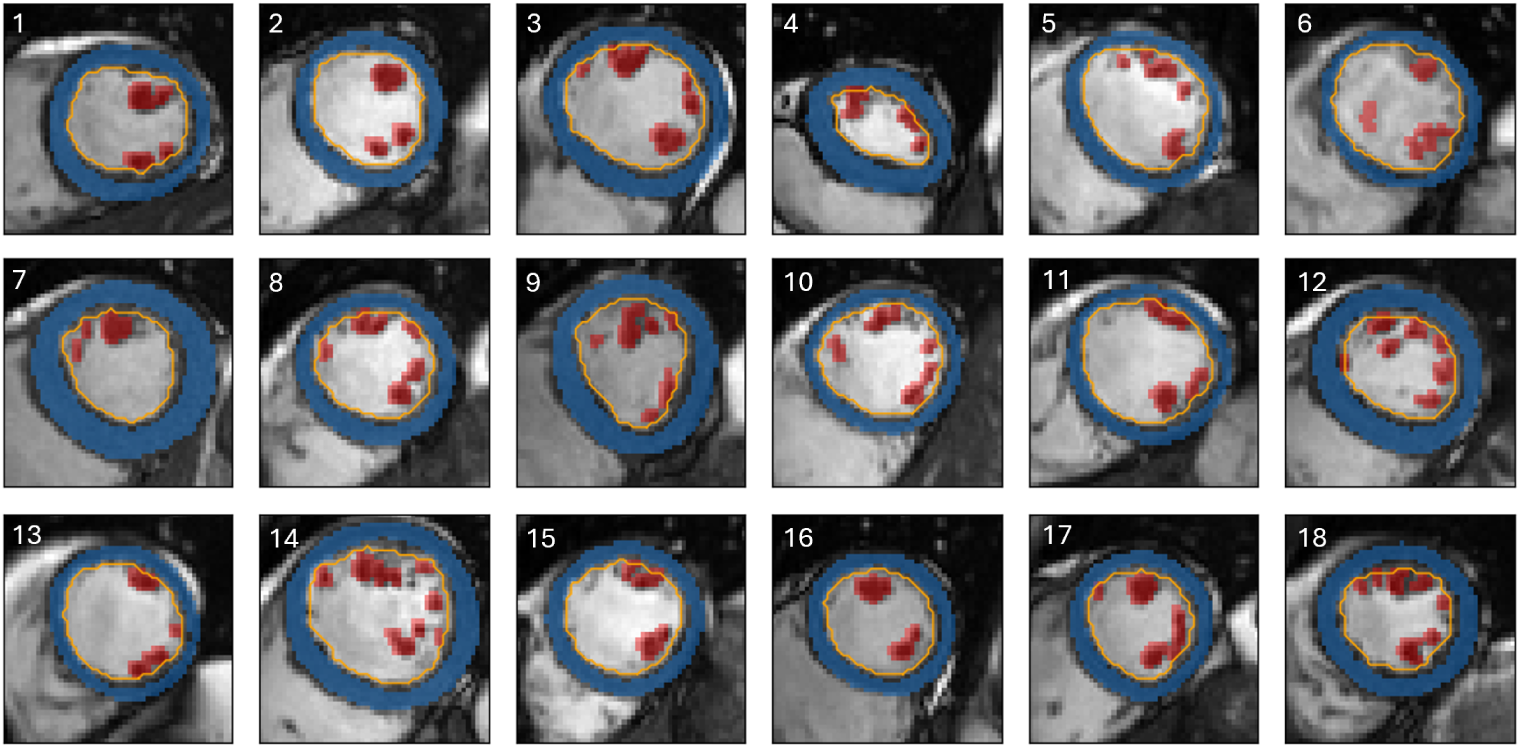
Segmentation results of the fd group show adequate quality. pm segmentations are indicated in **red**, myocardial segmentations in **blue**, and blood pool segmentations by **orange outlines**. cmr scans are reproduced by kind permission of UK Biobank ©.

Additionally, the robustness of the slice selection is assessed by gauging the deviation of the pm cross-sectional area measurement of the selected slice with adjacent slices.

The T frame is selected to depict the ED, where the left ventricle is the most dilated. This ensures the optimal differentiation of the PMs from each other and minimizes contact with the myocardium, which can lead to confusion with trabeculae. This is consistent with the measurements throughout the time points depicted in Figure 3 along the horizontal axis: Systolic time points (approximately T frames 10 to 25) of the heatmaps show smaller pm cross-sectional areas, as the left ventricular myocardium is constricted. On the other hand, diastolic time points (approximately T frames 40 to 10) depict the PMs fully. Additionally, the pm area measured in the selected T frame (indicated by the black outline) is largely consistent across multiple adjacent T frames, implying that some level of inaccuracy in the identification of the ed time point can be tolerated without risking substantial errors of measurement.

**Figure 3:**
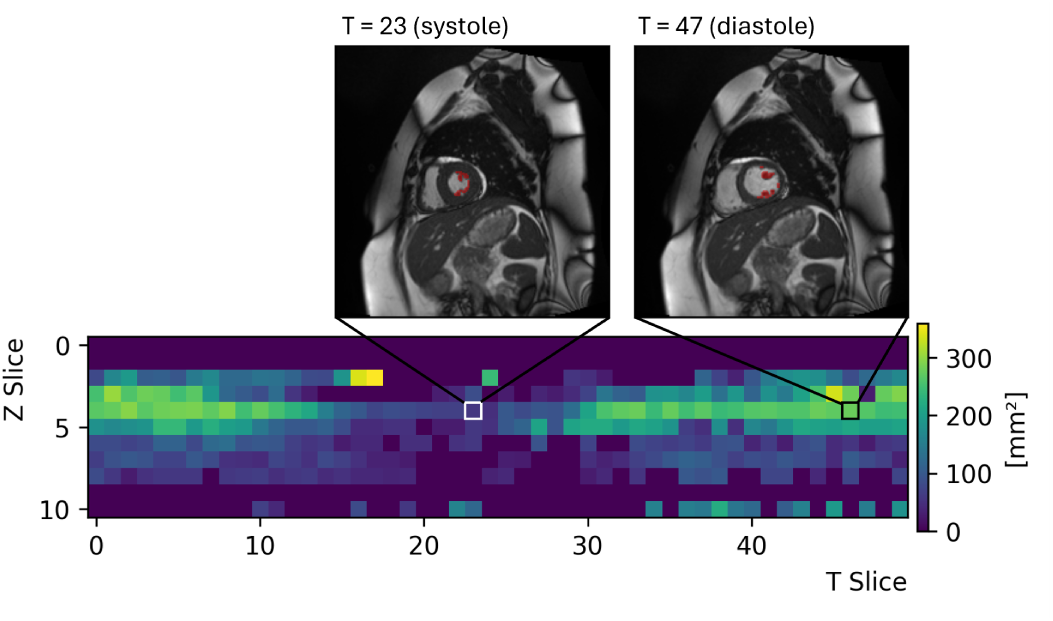
pm area is robust across T frames and reaches its maximum in the selected Z slice. Shown is a representative heatmap depicting pm cross-sectional area across all Z slices and T frames for the assessment of the slice selection robustness. The slice selected by the pipeline is indicated by a **black outline**. In addition, the cmr scan corresponding to the selected slice **(upper right)**, as well as one representative systolic time point **(upper left)**, are shown. The pm segmentation is overlayed in **red**. All other heatmaps of the fd group are depicted in Appendix B. cmr scans are reproduced by kind permission of UK Biobank ©.

Furthermore, the fifth Z slice (index 4) in each image is selected for the measurements. In this plane of the left ventricle, the blood pool is about at its largest and the PMs are typically clearly visible. This is also observed in Figure 3 in the center of the vertical axis. In more apical or more basal planes of the myocardium, the PMs may not be visible as clearly or at all, leading to lower pm cross-sectional areas. Because of this, even directly adjacent Z slices may yield very different measurements. The selected slice (indicated by the black outline) exhibits the highest measurements out of all Z slices in almost all cases. Thus, the selection of the Z slice is also deemed sufficiently consistent.

### 3.3 Control Group Selection

To evaluate the diagnostic potential of the biomarkers for FD, comparison with a cardiac-healthy control is required. For this, a control pool is created on the ukb RAP by excluding participants with cardiac diagnoses or self-reported heart problems. Exact exclusion criteria of the control pool are listed in Appendix C. This control pool is passed to the pipeline, which selects a specified number of trait-matched participants for each fd case. The pipeline can consider any number of traits available on the UK Biobank. Here, we select five control group participants with the same age and sex for each fd case. If not enough suitable controls are found in the control pool, a control with the next closest available age is selected instead. This approach yields a cardiac-healthy control group consisting of 90 participants, with a mean age of 63.92 years (age range 47 to 78). For comparison, the fd group exhibits a mean age of 64.00 years with the same age range.

Similarly, a second control group with a history of heart attacks is created to assess the specificity of the biomarkers. Criteria for inclusion into the pool are listed in Appendix D. Once again, five sex- and age-matched controls are selected for each fd group participant, yielding a heart attack control group consisting of 90 participants with a mean age of 64.02 years (age range 47 to 78).

### 3.4 Biomarker Characterization

#### 3.4.1 Diagnostic Relevance

First, the value of the cardiac metrics as diagnostic biomarkers is evaluated. Comparison of the fd group with the cardiac-healthy control group (Figure 4a, Table 3) allows for initial information about which markers are the most promising in this regard. The two lvh metrics, the myocardial cross-sectional area (ma) and the maxi-mum myocardial wall thickness (mwt), exhibit the largest deviations of the fd group from the controls and the lowest *p*-values. In contrast, the pmh metrics show smaller divergences from the control. The diagnostic performance is further assessed by plotting the roc curves (Figure 4b). Again, the two lvh metrics perform best out of the selected set, achieving the highest levels of sensitivity and specificity, as evidenced by their auc values of 0.69 and 0.66 for the mwt and the MA, respectively. As expected, these auc values are not high enough to qualify them as stand-alone markers, but they do suggest informative content that could be relevant in the diagnostic process, when combined with additional factors.

**Figure 4:**
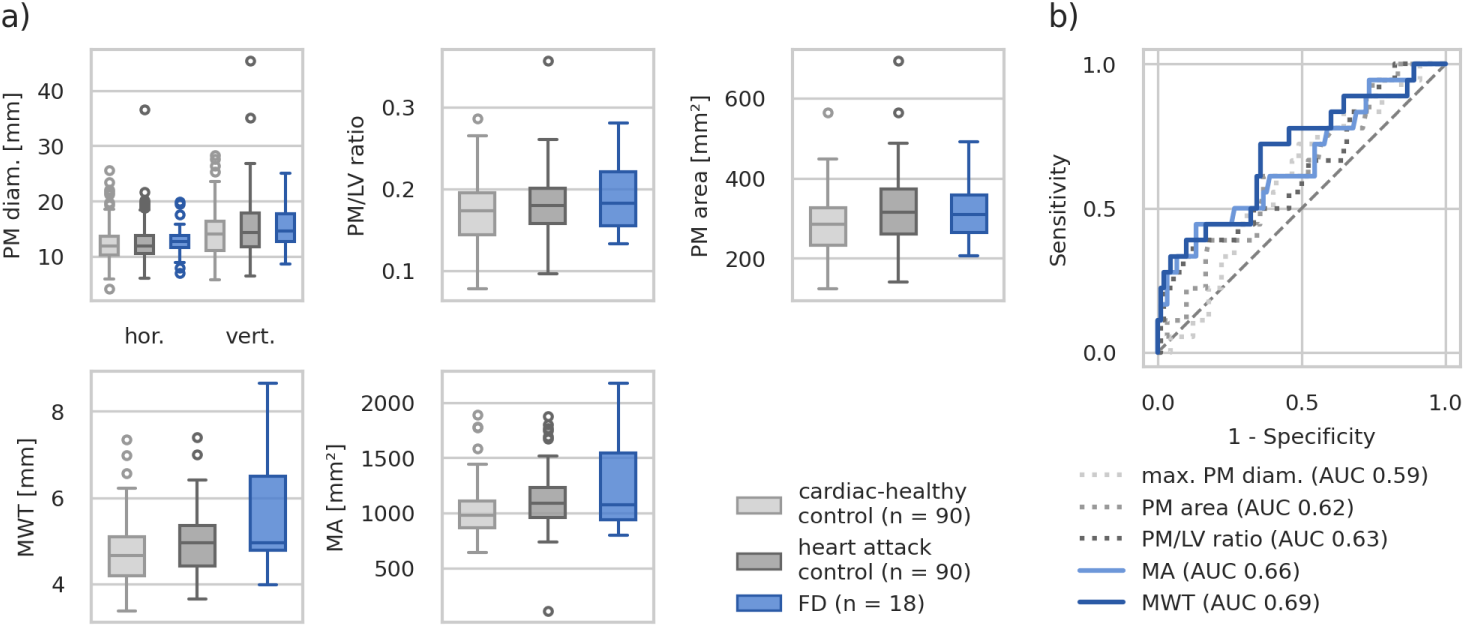
lvh metrics achieve the largest difference from the cardiac-healthy control and the highest roc AUCs. Subfigure a: Comparison of the analyzed cardiac metrics measured in the fd group **(right, blue)** with the sex- and age-matched cardiac-healthy **(left, light gray)** and heart attack control groups **(middle, dark gray)**. **Subfigure b:** roc curves of the cardiac metrics, calculated using the fd group as positives, and the cardiac-healthy control group as negatives.

**Table 3:** Statistical comparison of the cardiac metrics in the fd group and the cardiac-healthy control group.

|  | *p*-value | Cohen's *d* | *MD* |
| --- | --- | --- | --- |
| FD group compared to cardiac-healthy and heart attack control groups. |  |  |  |
| Horizontal PM diameters | 0.2248 | 0.10 | 0.39 mm |
| Vertical PM diameters | 0.0927 | 0.22 | 1.05 mm |
| PM/LV ratio | 0.0887 | 0.48 | 0.02 |
| PM cross-sectional area | 0.1163 | 0.36 | 33.87 mm<sup>2</sup> |
| MA | 0.0331 | **0.63** | 234.43 mm<sup>2</sup> |
| MWT | 0.0102 | **0.75** | 0.91 mm |
| FD group compared to heart attack control |  |  |  |
| Horizontal PM diameters | 0.3059 | 0.05 | 0.19 mm |
| Vertical PM diameters | 0.6123 | 0.00 | 0.03 mm |
| PM/LV ratio | 0.4263 | 0.26 | 0.01 |
| PM cross-sectional area | 0.9310 | 0.01 | 1.23 mm<sup>2</sup> |
| MA | 0.5175 | 0.35 | 141.23 mm<sup>2</sup> |
| MWT | 0.1202 | **0.57** | 0.70 mm |

Cohen's *d* values over 0.5 suggest medium or high effect sizes and are indicated by **bold** font. There are no *p*-values below the Bonferroni-corrected level of significance $\alpha_c = 0.0083$ .

To gauge the specificity of the markers to FD, comparison with a control group with a history of heart attacks is performed (Figure 4a, Table 3). Overall, the measurements are slightly higher in the fd group compared to the heart attack control group throughout most metrics, although all metrics clearly lack statistical significance or effect sizes large enough to be observable in a clinical setting. One notable exception could be the MWT, where the difference between the two groups—although not statistically significant—shows a medium to large effect size, as indicated by a Cohen’s *d* of 0.57.

To account for the heterogeneous fd presentation between sexes, the metrics are also analyzed after sex-stratification (Figure 5, Table 4). This reveals much more severe lvh in male fd cases compared to female fd patients and to male controls. The pmh metrics seem to be robust thoughout both subpopulations, based on our dataset.

**Figure 5:**
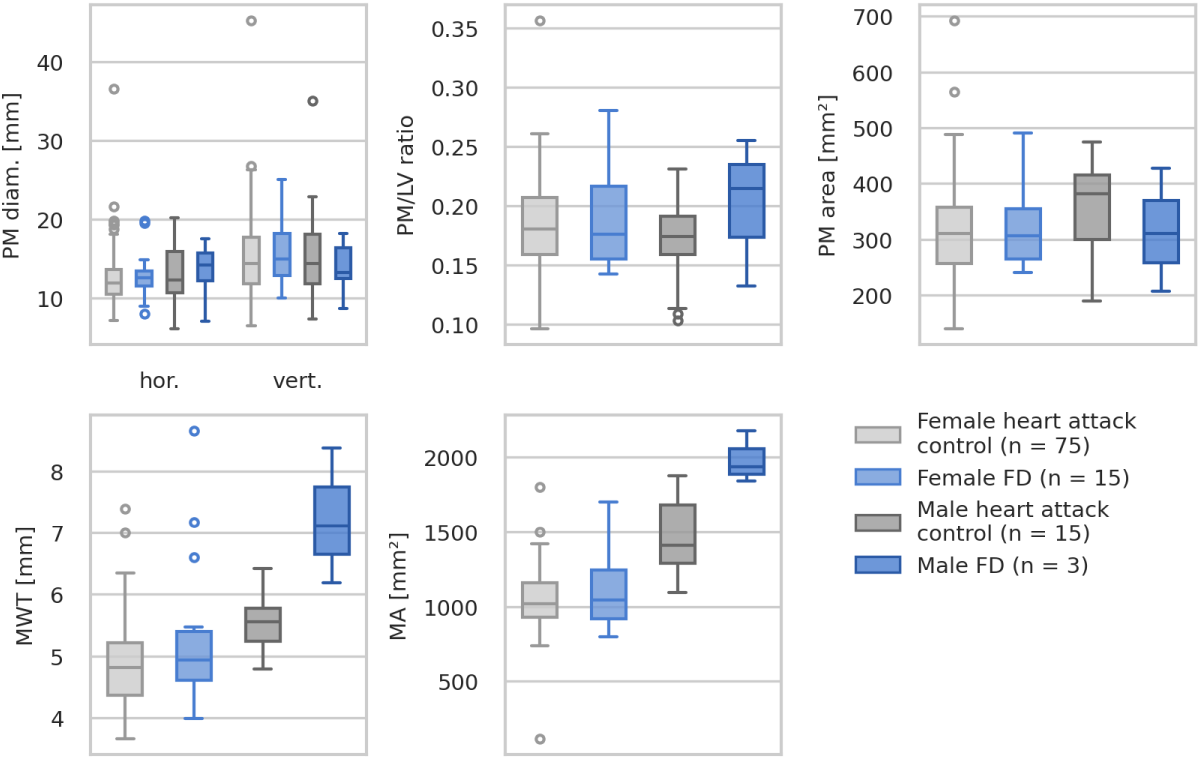
Male fd patients exhibit more severe lvh compared to female fd patients and cardiac-healthy controls. Shown is a comparison of the cardiac metrics measured in the female **(light colors)** and male **(dark colors)** subpopulations of the fd group **(blue)** and the corresponding sex- and age-matched cardiac-healthy control groups **(gray)**. Corresponding statistics are listed in Table 4.

**Table 4:**
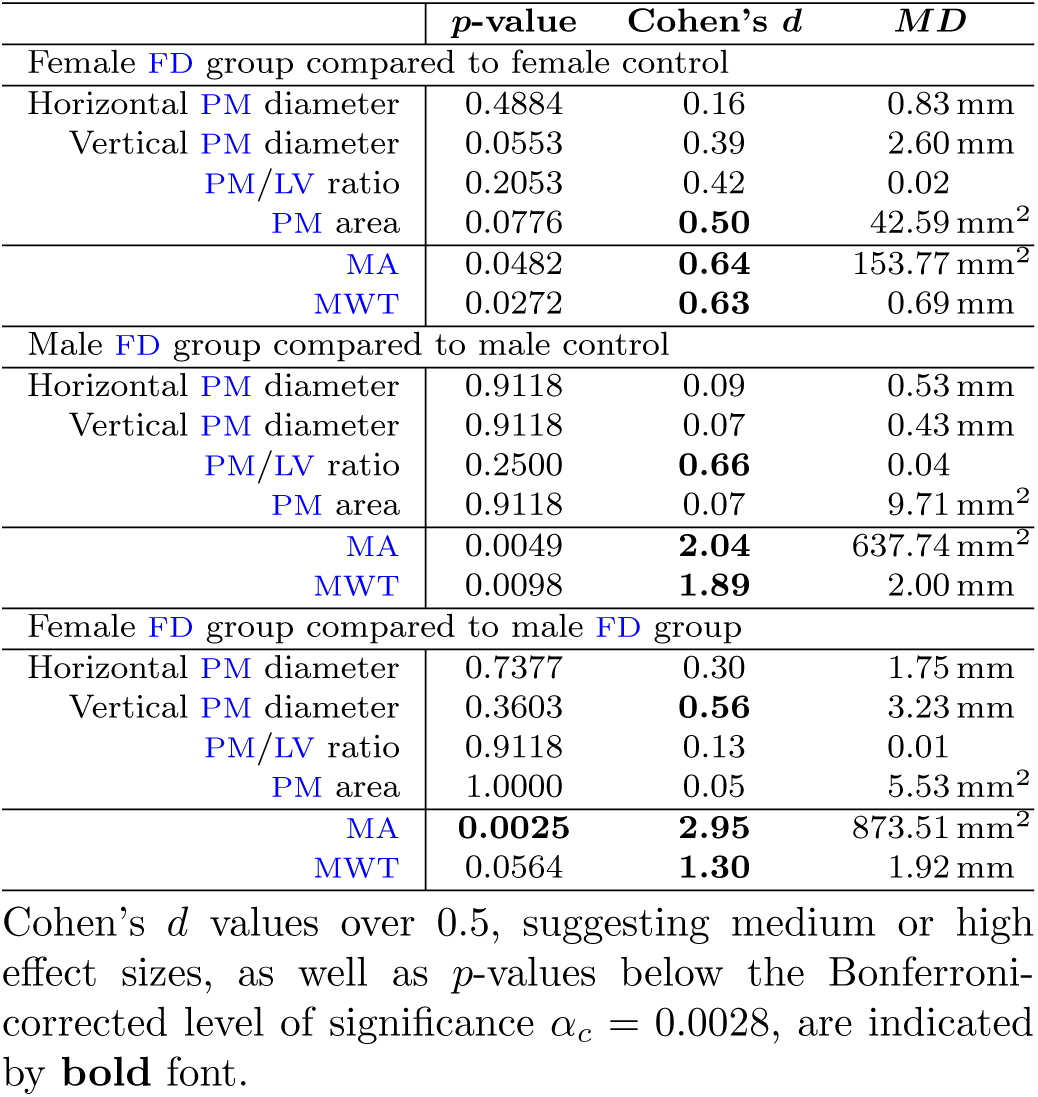
Statistical comparison of the cardiac metrics in the sex-stratified fd group and the cor-responding cardiac-healthy controls.

|  | *p*-value | Cohen’s *d* | *MD* |
| --- | --- | --- | --- |
| Female FD group compared to female control |  |  |  |
| Horizontal PM diameter | 0.4884 | 0.16 | 0.83 mm |
| Vertical PM diameter | 0.0553 | 0.39 | 2.60 mm |
| PM/LV ratio | 0.2053 | 0.42 | 0.02 |
| PM area | 0.0776 | **0.50** | 42.59 mm<sup>2</sup> |
| MA | 0.0482 | **0.64** | 153.77 mm<sup>2</sup> |
| MWT | 0.0272 | **0.63** | 0.69 mm |
| Male FD group compared to male control |  |  |  |
| Horizontal PM diameter | 0.9118 | 0.09 | 0.53 mm |
| Vertical PM diameter | 0.9118 | 0.07 | 0.43 mm |
| PM/LV ratio | 0.2500 | **0.66** | 0.04 |
| PM area | 0.9118 | 0.07 | 9.71 mm<sup>2</sup> |
| MA | 0.0049 | **2.04** | 637.74 mm<sup>2</sup> |
| MWT | 0.0098 | **1.89** | 2.00 mm |
| Female FD group compared to male FD group |  |  |  |
| Horizontal PM diameter | 0.7377 | 0.30 | 1.75 mm |
| Vertical PM diameter | 0.3603 | **0.56** | 3.23 mm |
| PM/LV ratio | 0.9118 | 0.13 | 0.01 |
| PM area | 1.0000 | 0.05 | 5.53 mm<sup>2</sup> |
| MA | **0.0025** | **2.95** | 873.51 mm<sup>2</sup> |
| MWT | 0.0564 | **1.30** | 1.92 mm |

Cohen’s *d* values over 0.5, suggesting medium or high effect sizes, as well as *p*-values below the Bonferroni-corrected level of significance $\alpha_c = 0.0028$ , are indicated by **bold** font.

Based on these analyses, the mwt emerges as the most promising candidate for a diagnostic biomarker. The robustness regarding non-concentric lvh enabled by the ma metric does not seem beneficial. In contrast, pmh is not found to be useful for diagnostic purposes based on the available data.

#### 3.4.2 Prognostic Value

Next, the metrics are evaluated for their prognostic value. For this, direct correlation of the metrics with disease progression or severity would be ideal. However, disease progression is not easily quantifyable given the data available on the UK Biobank, as there are no suitable health-related records or repeat imaging data of any fd cases at this time. Instead, we exploit the typical disease progression of FD, employing age as a proxy for disease progression. Correlation of the cardiac metrics with age shows a significant and strong association of the PM/LV ratio in the fd group but not in the cardiac-healthy control group (Figure 6). This could suggest a relevant role of the PM/LV ratio as a prognostic marker, as it likely increases with disease progression and severity.

**Figure 6:**
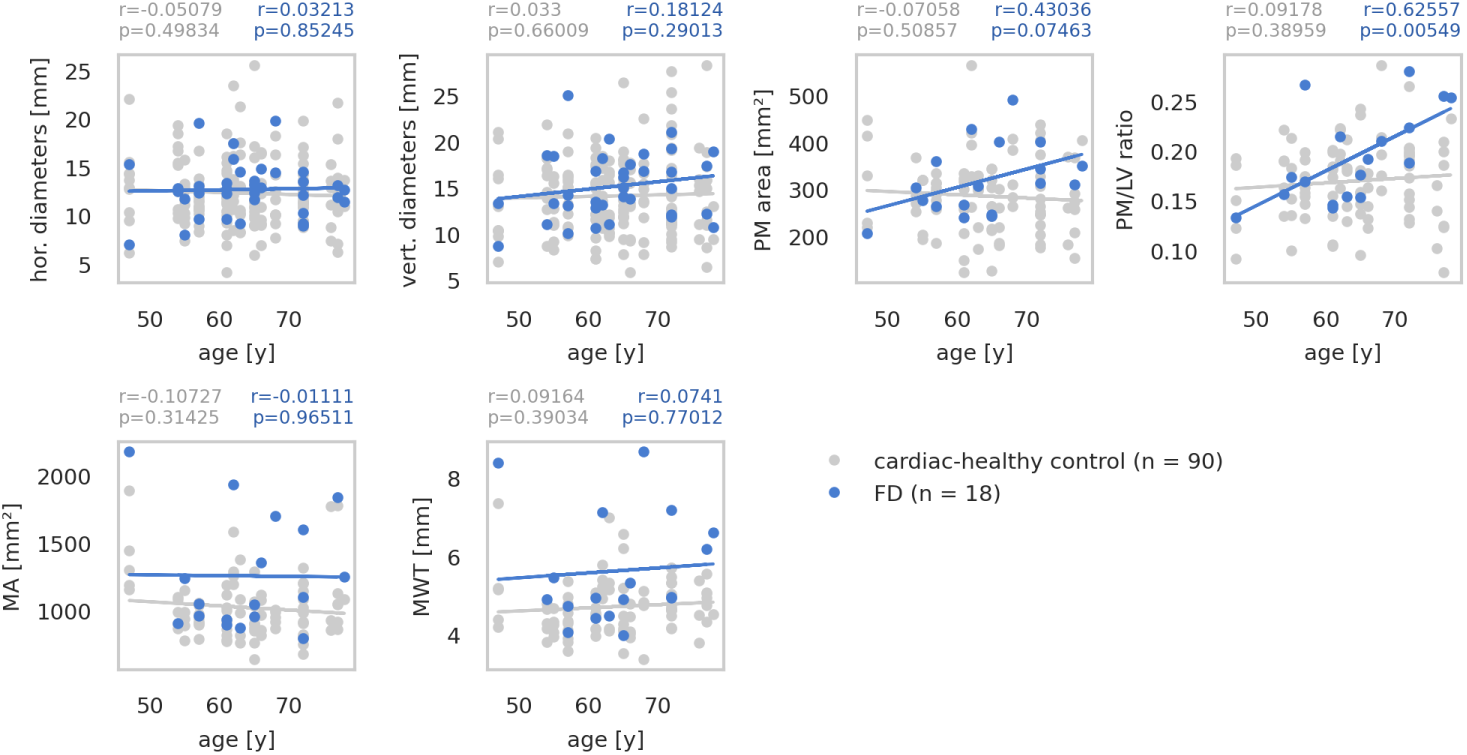
The PM/LV ratio increases with age in the fd group, contrasting with the other metrics. Shown are scatter plots and corresponding linear fitted curves depicting the age-correlation of each of the cardiac metrics in the fd group **(blue)** and the cardiac-healthy control group **(gray)**. The Pearson correlation coefficient *r* and the corresponding *p*-value are noted above each plot. The Bonferroni-corrected level of significance *α_c_* is 0.0083.

Cianciulli et al. [4] presented some evidence for PMH, specifically in the form of the pm diameters, as a predictive marker for LVH. To assess whether this is the case in our cohort, we first calculated pairwise correlation between the pmh and lvh metrics to identify relationships that might indicate possible predictive roles. In contrast to defining fixed thresholds differentiating the binary presence or absence of pmh or LVH, this approach preserves the continuity of the data. The pair correlation shows that some pairs, such as the pm cross-sectional area and the MWT, exhibit differing correlation patterns in the fd group and the cardiac-healthy control group, although neither is significant (Figure 7). While this could suggest a predictive relationship between the metrics, it cannot elucidate its direction. Notably, no significant correlation between the pm diameters and any other metric is observed.

**Figure 7:**
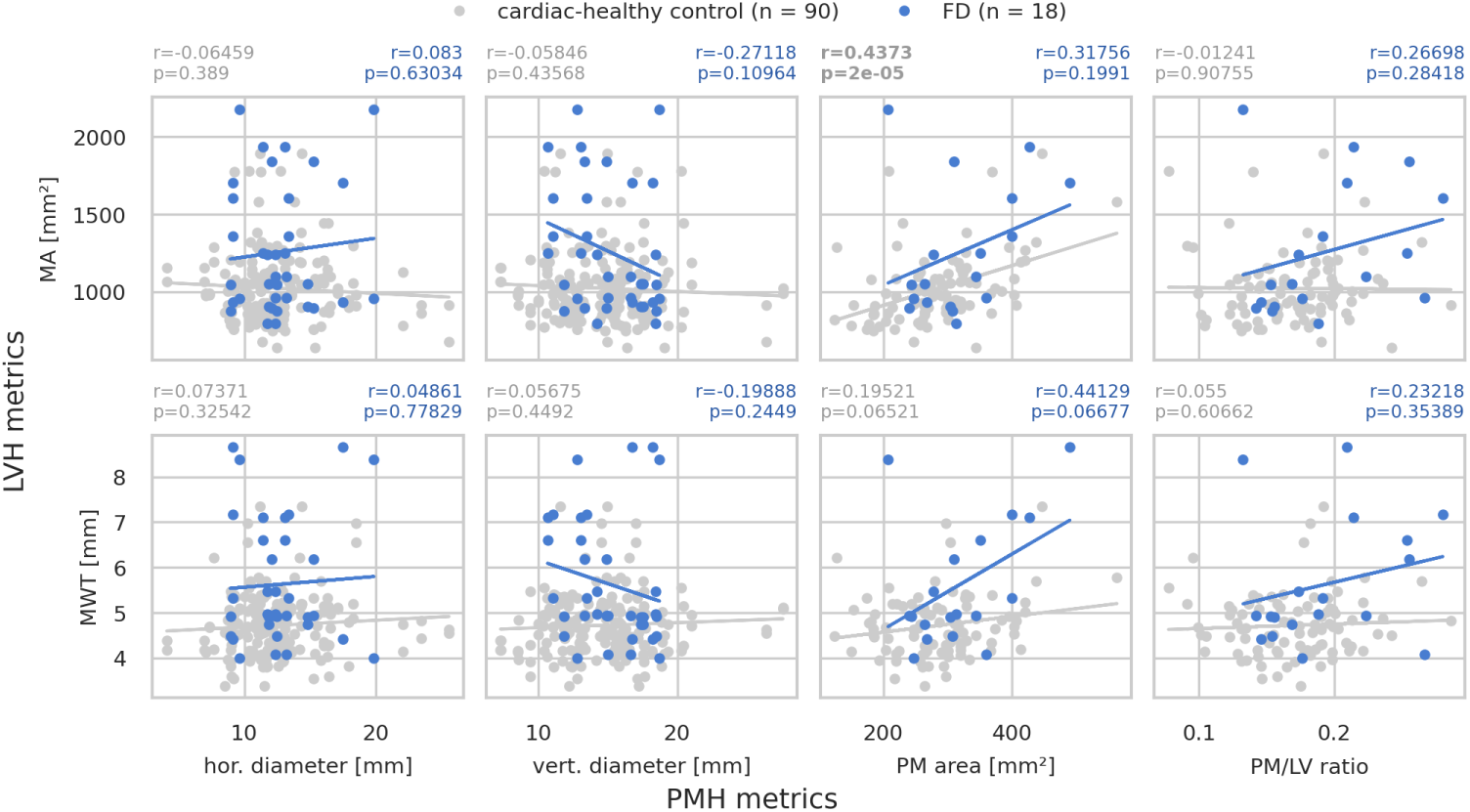
The relationship between the pm cross-sectional area and the mwt differs from the cardiac-healthy control. Shown are scatter plots and corresponding linear fitted curves depicting the correlation of each of the pmh metrics with each of the two lvh metrics in the fd group **(blue)** and the cardiac-healthy control group **(gray)**. The Pearson correlation coefficient *r* and the corresponding *p*-value are noted above each plot. The Bonferroni-corrected level of significance *α_c_* is 0.0031.

**Figure 8:**
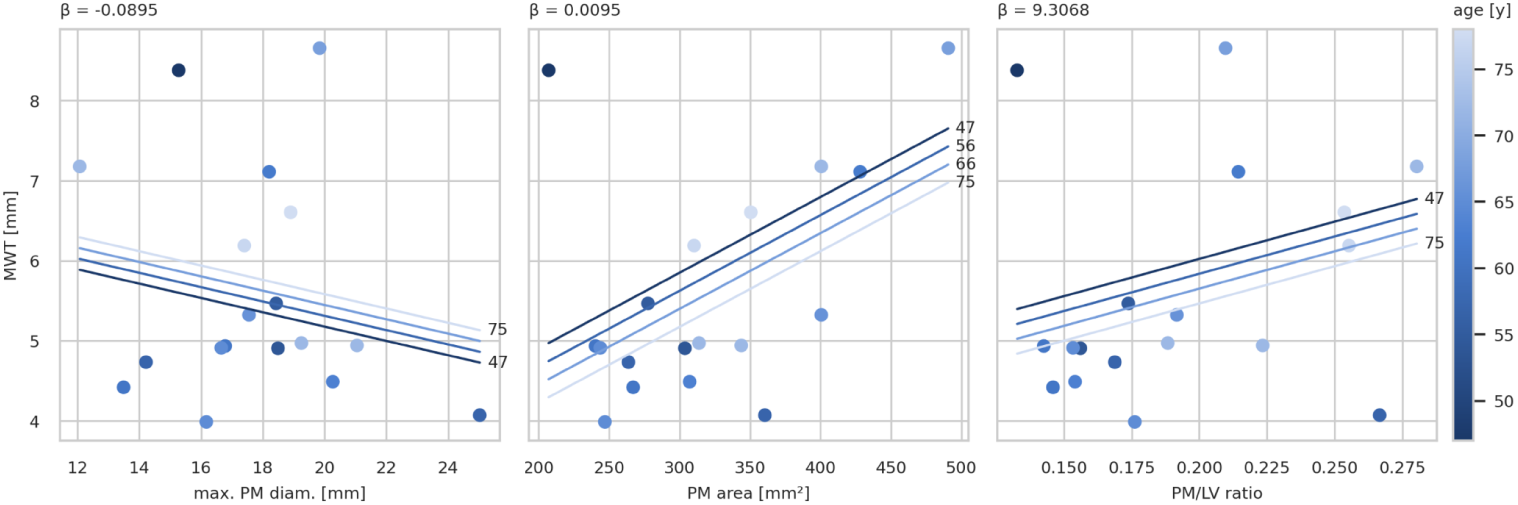
The pm cross-sectional area shows an age-independent effect on the lvh, suggesting possible predictive power. Multiple regression of the mwt as the dependent variable with age and each of the three pmh metrics, the maximum pm diameter **(left)**, the pm area **(middle)**, and the PM/LV ratio **(right)**, as the independent variables. Regression planes are simplified as sets of lines for ease of visualization. Multiple regression analyses with switched dependent and independent variables are shown in Appendix E.

Assuming that both the pmh metrics and the lvh metrics increase with age and disease progression in the general fd population, their relationship could also arise from their shared age-correlation. To gauge the actual effect of the pmh metrics on the LVH, ols multiple regression is performed with the mwt as the dependent variable and the pmh metrics and age as the two predictors. This is repeated for all three pmh metrics.

Regardless of the tested pmh metric, the slope *β_MW T_*, which describes the effect of age on the mwt when holding the pmh metric constant, is close to zero and clearly non-significant. Importantly, this may differ in younger cohorts.

Out of the three pmh metrics, the pm cross-sectional area exhibits the largest age-independent effect on the MWT, followed by the PM/LV ratio. These effects are likely large enough to be observable in a clinical setting. However, the *p*-value of the effect of the PM/LV ratio of 0.36, indicating that this effect lacks statistical robustness, making it unreliable as a predictive marker. The *p*-value corresponding to the effect of the pm cross-sectional area is closer to the level of significance. For the pm diameters, the maximum value out of the four diameters measured from each participant is tested following the approach of Cianciulli et al. [4]. In this cohort, the pm diameters do not exhibit a positive effect on the MWT, which stands in contrast to previous hypotheses [4].

## 4 Discussion

Building on previous research, we have analyzed the diagnostic and prognostic value of previously proposed cardiac biomarkers in FD. Some of the metrics, namely the pm diameters proposed by Cianciulli et al. [4] and the PM/LV ratio proposed by Niemann et al. [8], are originally established in echocardiography. We expanded on this by applying them to short-axis cmr imaging in a new UK-based cohort. Our fully automated image analysis pipeline enables time-efficient measurement of the metrics while minimizing inter- and intraobserver variability. Using this pipeline, we performed a comparative analysis of the clinical value of the metrics.

### 4.1 Left Ventricular Hypertrophy Exhibits Diagnostic Value in Fabry Disease

The comparative analysis of the proposed cardiac metrics confirms the known diagnostic value of lvh for FD. Out of the assessed metrics, the two lvh metrics—the maximum myocardial wall thickness (mwt) and the myocardial cross-sectional area (ma)—exhibit the largest deviations from the cardiac-healthy control group and achieve the highest roc AUCs, outperforming the pmh metrics. While neither metric performs well enough to be useful as a stand-alone marker, as expected, they carry diagnostic information that may be useful when combined with additional factors.

Further analyses reveal some evidence that the mwt might be more specific for fd when compared to a heart attack control, although this effect may not be reliable for clinical application. The increased robustness of the ma regarding non-concentric lvh does not seem to benefit its diagnostic performance. Throughout the analyses, we did not find evidence for a prognostic potential of the lvh markers. In summary, these results match well with previous research [2] and current clinical practice relying on the diagnostic value of lvh [9].

### 4.2 Papillary Muscle Hypertrophy Shows Evidence for Prognostic Role

In contrast, the pmh metrics are not found to be useful for diagnostic purposes. Initial evidence, however, suggests prognostic potential. The PM/LV ratio stands out among the metrics for its significant increase with age, a proxy for progression of FD. Based on this, the PM/LV ratio might be useful to gauge the disease severity in FD.

Furthermore, we investigate the possible predictive power of the pmh metrics regarding the LVH, as proposed by Cianciulli et al. [4]. Here, we found that the pm cross-sectional area—and to a lower degree the PM/LV ratio—show observable effects on the mwt when correcting for age as a confounding factor. Notably, the pm di-meters do not exhibit an effect on the MWT, which stands in contrast to previous interpretations [4]. This implies a predictive relationship between the pmh and the LVH, although further analyses are necessary for a clear interpretation.

### 4.3 Limitations

Importantly, the available fd cohort is small (*n* = 18) and has a limited age range, with the youngest participant being aged 47 at the time of imaging. Within this cohort’s age range, cardiac involvement is already expected to be well established in most fd patients, limiting conclusions regarding early onset of these markers and the prognostic or predictive power of the analyzed metrics. There is no information available on FD-specific therapeutic intervention. Thus, there is a clear need for a younger and larger cohort, where we expect repeat analyses to yield clearer results, with increased stability of the effect of the pmh metrics on MWT. It is also plausible for additional age-correlation patterns or potential predictive roles to be revealed in a younger cohort, particularly regarding the pmh metrics, as previous literature hypothesizes early onset of these markers [4]. The UK Biobank’s extensive data collection and imaging protocols also introduce a bias towards participants with less severe disease presentations, as participants with debilitating symptoms are less likely to volunteer for this procedure. However, collecting high-quality and comparable data of patients with rare and under-or misdiagnosed diseases such as fd remains a central challenge.

## 5 Conclusion

Our analyses confirm the known role of lvh as a diagnostic marker [2, 3, 9]. We found that the MWT, which mimics the method established for diagnosing lvh in a clinical setting, performs slightly better as a diagnostic marker in our cohort than the MA, which takes the entire left ventricular myocardium into account. Additionally, we found some initial evidence for PMH, especially the PM/LV ratio and the pm cross-sectional area, to carry relevant prognostic information. For our analyses, we have established a fully automated image processing pipeline that could serve as a valuable foundation for further research on imaging-based cardiac biomarkers.

## Data Availability

All data analyzed in the present study are provided by the UK Biobank.

## Abbreviations

AUC: area under the curve
CMR: cardiac magnetic resonance imaging
DICOM: Digital Imaging and Communications in Medicine
ED: end-diastole
FD: Fabry disease
FOV: field of view
FPR: false positive rate
ICD-10: International Classification of Diseases Version 10
LV: left ventricle
LVH: left ventricular hypertrophy
MA: myocardial cross-sectional area
MRI: magnetic resonance imaging
MWT: maximum myocardial wall thickness
OLS: ordinary least squares
PM: papillary muscle
PMH: papillary muscle hypertrophy
ROC: receiver operating characteristic
TPR: true positive rate
TRUFI: true fast imaging with steady-state free precession
UKB RAP: UK Biobank Research Analysis Platform

## Acknowledgements

This research has been conducted using the UK Biobank Resource under Application Number 541112. Grammarly and DeepL were used to improve readability and wording of the manuscript.

## Declarations

### Ethics Approval

The UK Biobank has received ethical approval from the North West Multi-Center Research Ethics Committee (11/NW/0382). This research project was approved by the UK Biobank under the application number 541112.

### Consent for Publication

Not applicable.

### Data Availability

All raw data is available on the UK Biobank. All measurements and statistical analyses are available on GitHub (BioMeDS/Fabry Cardiac Quantification).

### Materials Availability

Not applicable.

### Code Availability

- Project name: Fabry Cardiac Quantification
- Project home page: https://github.com/BioMeDS/Fabry Cardiac Quantification
- Archived version: 10.5281/zenodo.19818354
- Operating system(s): designed for use on the UKB-RAP, but functions platform-independently
- Programming language: Python
- Other requirements: `ukbb_cardiac` [11]
- License: MIT
- Any restrictions to use by non-academics: none

### Author Contribution

M. Schüttler developed the image processing pipeline, performed the presented analyses, and wrote the main manuscript text. J. Witte performed the genetic identification of the fd participants. M. Schindehütte and P. Nordbeck provided clinical insight. M. Schindehütte and M. Ankenbrand supervised the project. All authors revised and contributed to the final manuscript.

## Appendix A Genetic Variants Observed in the fd Group

The following *GLA* gene variants are observed in the fd CMR imaging group: N215S, R363C, R356Q, R112H, and I198T, as well as non-exonic mutations.

## Appendix B Full Results of the Slice Selection Robusness Assessment

**Figure B1:**
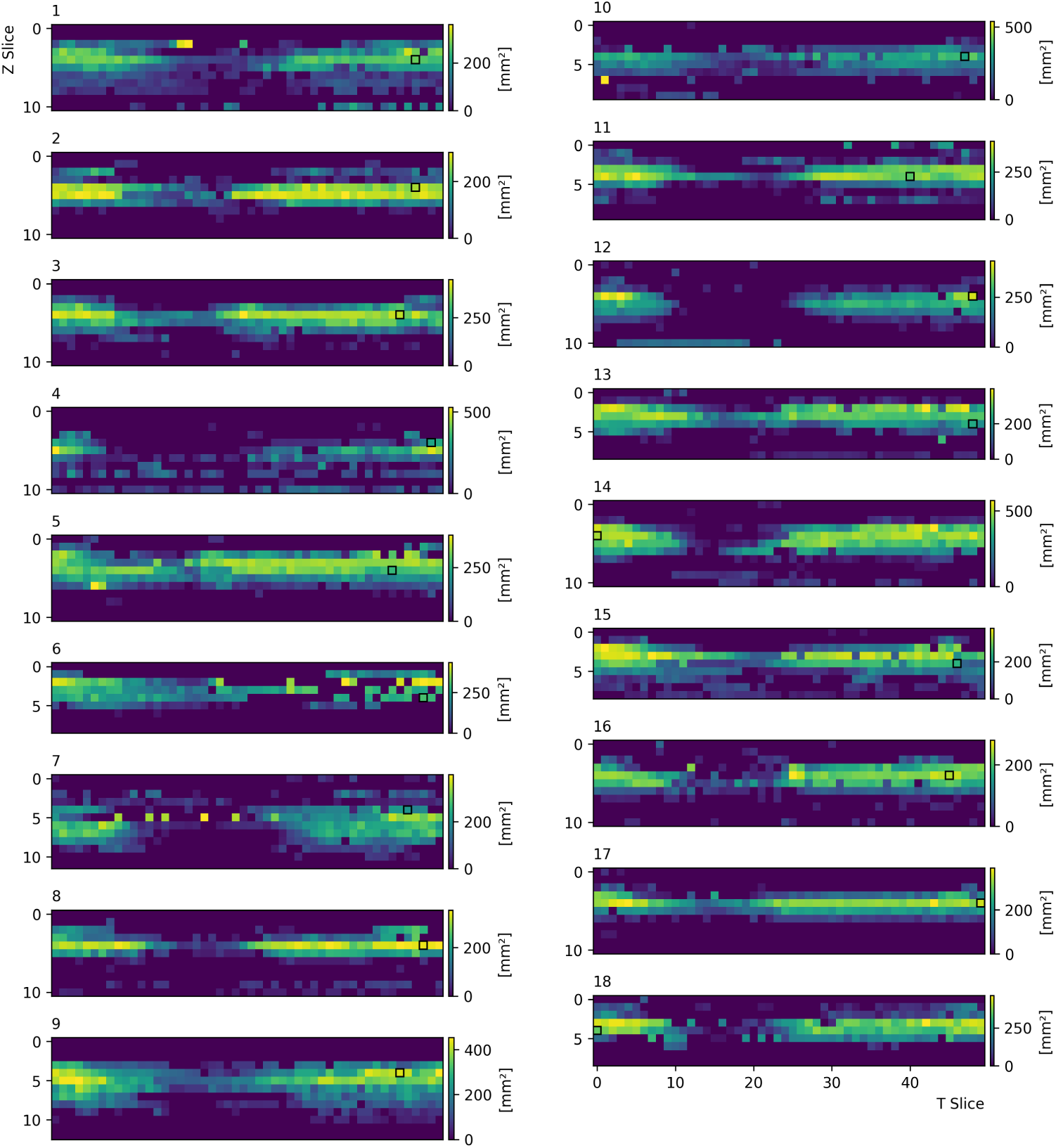
Heatmaps depicting the pm cross-sectional area across all Z slices and T frames of the eighteen fd group images for the assessment of the slice selection robustness. The slices selected by the pipeline are indicated by a black outline. Values within the white outline are taken into account for the calculation of the standard deviation *SD*.

## Appendix C Definition of the Cardiac-Healthy Control Pool

The cardiac-healthy control pool cohort was created by exclusion of several factors. These are:

- Identification as Fabry participant,
- Diagnoses of cardiac diseases (ICD-10, Data-Field 41270):

– C38: Malignant neoplasm of heart, mediastinum and pleura,
– I80-89: Diseases of veins, lymphatic vessels and lymph nodes, not elsewhere classified,
– I95: Hypotension,
– Q20-Q28: Congenital malformations of the circulatory system,
– Q87: Other specified congenital malformation syndromes affecting multiple systems,
– R00: Abnormalities of heart beat,
– R01: Cardiac murmurs and other cardiac sounds,
– R03: Abnormal blood-pressure reading, without diagnosis,
– T82: Complications of cardiac and vascular prosthetic devices, implants and grafts,
– T86.2: Heart transplant failure and rejection,
– T86.3: Heart-lung transplant failure and rejection.
- Disclosure of any diagnosis regarding vascular or heart problems during a digital assessment performed on the same day as the short axis cmr scan acquisition (Data-Field 6150),
- Disclosure of current heart issues during a digital follow-up assessment (Data-Field 28675),
- Lack of available short-axis cine cmr imaging data.

This control pool comprises 4506 individuals (as of August 11, 2025).

## Appendix D Definition of the Heart Attack Control Pool

The heart attack control pool was created by including participants with history of heart attacks:

- Diagnoses of heart attack (ICD-10, Data-Field 41270):

– I21: Acute myocardial infarction,
– I22: Subsequent myocardial infarction.

Additionally, participants with these factors are excluded from the control pool:

- Identification as Fabry participant,
- Lack of available short-axis cine cmr imaging data.

This control pool comprises 1849 individuals (as of August 11, 2025).

## Appendix E Supplementary Results of the Multiple Regression Analysis

**Figure E1:**
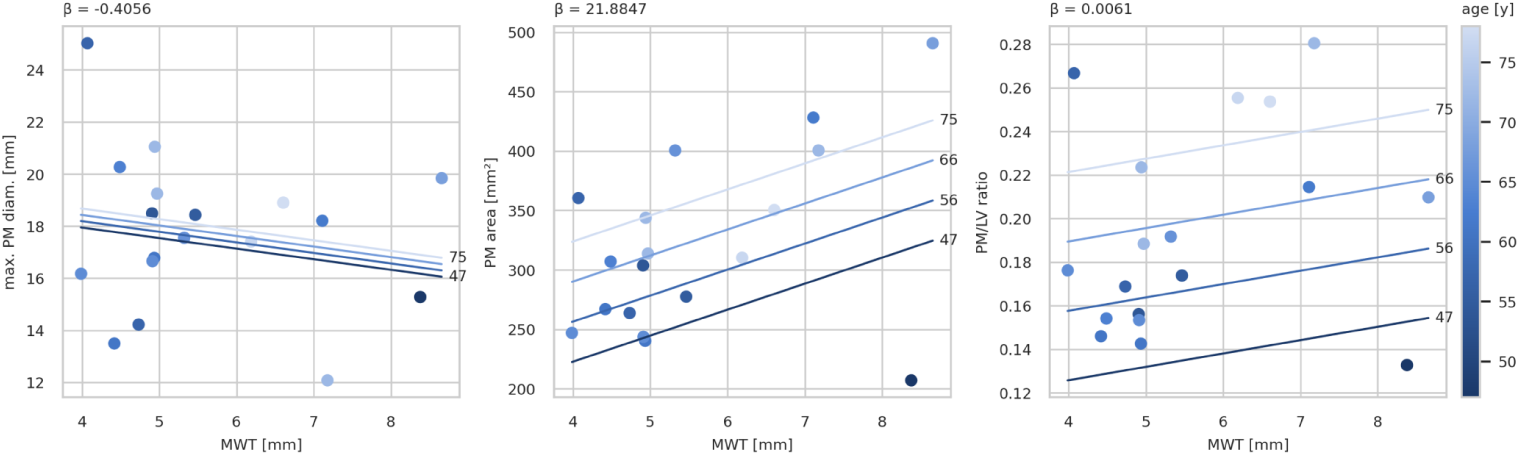
Multiple regression of each of the three pmh metrics as the dependent variable with age and mwt as the independent variables.

